# Assessing antimicrobial resistance, utilization and stewardship in Yemen: An exploratory mixed-methods study

**DOI:** 10.1101/2021.01.27.21250628

**Authors:** ESF Orubu, A Najwa, C Ching, S Bu Jawdeh, J Anderson, R Sheikh, F Hariri, H Basaleem, MH Zaman

## Abstract

Antimicrobial resistance (AMR), largely driven by irrational use of antimicrobials, is a global, multi-faceted problem calling for a complete understanding of all contributory factors for effective containment. In conflict settings, war-wounds and malnutrition can combine with existing social determinants to increase demand for antibiotics, compounding irrational use. In this study, we focus on Yemen, a low-income country with active conflict for the last five years, and analyze the current status of awareness and stewardship efforts regarding AMR. We performed a survey of prescribers/physicians and pharmacists to describe perceptions of AMR prevalence, antibiotic use practices and stewardship in Yemen, supported by a non-systematic scoping literature review and a key informant interview. Participants (96%, n=57) reported a perceived high AMR prevalence rate. Prescribers (74%, 20/27) reported pressure to prescribe broad-spectrum antibiotics. In the majority of cases (81%, 22/27), Antimicrobial Sensitivity Tests (AST) were not performed to inform antibiotic choice. The main barrier to AST was cost. Most pharmacists (67%, 18/27) sold antibiotics without prescriptions. Amoxicillin (including amoxicillin-clavulanate) was the most-commonly prescribed (63%, 17/27) or dispensed (82%, 22/27) antibiotic. AST was rated the least important solution to AMR in Yemen. While there was awareness of a high AMR rate, stewardship is poor in Yemen. We note that barriers to the use of AST could be addressed through the deployment of low-cost AST kits. Compulsory continuing education emphasizing the use of AST to guide prescribing and patients’ awareness programs could help avoid irrational use.

## Introduction

Antimicrobial resistance (AMR) – the ability of disease-causing microorganisms such as bacteria to withstand therapeutic doses of an antimicrobial agent, leading to treatment failure/death – is a multi-dimensional biosocial problem affecting countries worldwide and requires urgent measures aimed at containment.^1,2^ Low- and Middle-Income Countries (LMICs) are particularly projected to bear the greater impact of AMR.^1^ This may be even more so in humanitarian and armed conflict contexts.^3–5^

While a natural adaptive strategy, AMR has been accelerated by anthropogenic factors promoting inappropriate use, including non-evidence based prescribing, or empirical prescribing, self-medication, non-adherence to prescribed dosages, use of Substandard and Falsified (SF) medicines and non-therapeutic uses in livestock.^6–10^ Social determinants, including poor infection prevention and control practices, poor Water, Sanitation and Hygiene (WASH) infrastructure and lack of access to quality health services further exacerbate AMR, resulting in situations whereby antibiotics are used as substitutes for disease-preventive activities in some settings.^9,11,12^ Overall, more than 50% of all antibiotic prescriptions are inappropriate, calling for better Antimicrobial Stewardship (AMS) to preserve efficacy ^13^.

Specifically, in active conflicts, increased injuries associated with war wounds predisposing to AMR infections contribute additional contextual challenges.^4,14–18^ Some studies report or suggest a high carriage of AMR and transmission among refugees and the forcibly displaced fleeing conflicts in five World Health Organization (WHO) Eastern Mediterranean Region countries.^16,19^ Thus, conflicts aggregate multiple social determinants potentially promoting inappropriate antibiotic use and AMR.

Yemen, following upon upheavals arising from the Arab Spring in 2011 slipped into conflict in March 2015, and is now described as the world’s largest humanitarian crisis.^20^ In a population of 29.1 million in 2019, 24 million are in need of aid with about 3.3 million Internally Displaced Persons (IDPs). Access to healthcare, as well as to other services essential to life, remain limited. Since 2016, Yemen has been struggling with a cholera outbreak, leading to over 1.2 million cases as of January 2020; mostly in children under five^21^. Malnutrition in young children predisposing to infections is high.^22,23^ Several studies already suggest a high AMR burden.^24–30^ In 2018, for example, among patients admitted to the Médecins Sans Frontiers (MSF) hospital in Aden, more than 60% had a drug resistant infection.^31^ These conditions coincide with the current COVID-19 pandemic to further exacerbate the problem.^32^

Understanding and accounting for contextual factors impacting AMR, including individual, social and national-level dimensions, is necessary for the design and implementation of successful containment programs. Globally, the WHO’s Global Action Plan on AMR Containment, launched in 2015, articulates a strategic framework of actions. The follow-up 2017 Access, Watch, Reserve (AWaRe) classification of antibiotics aims to promote rational use at the community-level. Both lay emphasis on Antimicrobial Sensitivity Tests (AST) to guide antimicrobial prescribing and prescription-only-sale of antibiotics as two complementary AMS approaches. In general, surveillance data on AMR prevalence and antimicrobial consumption inform country-level measures to contain AMR.

There are surveillance gaps for Yemen that lead to a poor understanding of AMR drivers and containment efforts. There are no records on national AMR prevalence on several databases including the WHO’s GLASS, the PFIZER database and CDDEP resistance bank.^33^ There is also no national action plan on AMR. Conflict and a weak National Health Information System make it difficult to access reliable, up-to-date data.

The aim of this exploratory mixed-method study was to assess current awareness and perceptions of AMR and AMS status in Yemen. The objectives were to: 1. assess perceptions on AMR prevalence, 2. assess antibiotic prescribing and dispensing practices, and 3. evaluate AMS activities, with a view to both understanding current challenges with AMR and to provide evidence for policy interventions.

## Methods

Study design: The study employed a mixed-method study design. The mixed-method approach was judged appropriate to provide robust results; recognizing both the challenge of conducting research during conflicts in the middle of a pandemic and the need for accurate data. In designing, implementing and reporting this study, we adopted the mixed-method guidelines of Leech and Onwuegbuzie.^34^ It utilized a combination of a literature review, questionnaire survey and a focused key stakeholder interview.

### Literature review

To better understand the context and to evaluate our findings, we performed a targeted non-systematic literature review of grey, peer-reviewed and unpublished documents, reports and articles obtained from various sources. Documents selected *a priori* for evaluation included the National Essential Medicines List (EML). Peer-reviewed literature retrieved from a search on PubMed and Web of Science and reports identified from researchers’ experience in Yemen from 1999 were selected and evaluated, with a focus on the time period from 2015-2020. Articles related to AMR prevalence, prescribing and dispensing practices, antibiotic use and use of AST in Yemen, as well as other relevant policies including prescribing guidelines were selected and data was subsequently extracted. Thus, the literature review was iterative – performed before the survey to guide questionnaire development and after the survey to triangulate survey findings as well as support the key stakeholder’s interview to summarize and synthesis the current evidence.^34^

### Survey

#### Study participants

Study participants were physicians in private and public facilities that prescribe medicines and pharmacists in private pharmacies.

#### Sample size

We targeted a sample size of 40-60, comprising at least 30 physicians and 10 pharmacists, considered appropriate for a pilot or exploratory study.^35,36^

#### Study location

Facilities across Yemen, encompassing the “regional” north and south to account for the current administrative structures, comprising all five tiers of health units, health centers and hospitals (district, regional and central specialized). This dispersion allowed for a maximum variation sampling scheme to collect a wide range of perspectives.

Study instrument: Two different semi-structured questionnaires were used to collect data with tailored questions for healthcare workers/physicians and pharmacists (Appendix 1, Supplement). Questions were based on a rational construct to scope perceptions, practices and AMS activities in the relevant facility type. The construct included selected aspects of AMS with a focus on rational prescribing – the use of guides, antibiotic choice, prescribing (or consumption) patterns, the presence of Drug and Therapeutic Committees (DTC) and AST derived from WHO guidelines.^37,38^ The questionnaires contained four sections: (i) demographic information on both the clients of the facilities and participants; (ii) perceptions of the AMR burden and approaches aimed as solution; (iii) AMS; and (iii) practices related to antibiotic prescribing and dispensing, including the major antibiotics prescribed or dispensed. Questionnaires were made available in English and Arabic versions. Translations were provided by a proficient bilingual speaker (SB) and checked by a researcher (NA) who is also a proficient bilingual professional for accuracy, cultural suitability and content validity.

Survey administration and data collection: The questionnaires were distributed online for self-completion in August-September, 2020.

### Key informant interview

The interview was conducted with a public official in Yemen Republic, to triangulate information from the survey and literature review; both to understand public policies and to provide contextual information. Questions were formulated based on the survey results. The pertinent questions asked the key informant were: What factors would improve the uptake of AST, the average cost of one AST, and policies around prescription guides and AMR. To improve data quality, we purposively identified and interviewed a high-ranking official.

Using this politically-important case sampling strategy, we hoped to obtain accurate information on policies. The interview was conducted over the telephone using specific questions prepared in advance from survey findings.

### Analysis

The results were analyzed by descriptive statistics and presented as percentages. Survey responses with incompletely or incorrectly filled variables were screened out before data analysis. Interview findings are reported qualitatively as received.

### Ethics

Ethical clearance (REC-81-2020) was obtained from the Research Ethics Committee of the University of Aden – in line with similar studies – before questionnaires were distributed.^24^

## Results

The survey response rate was 41% (57/140). There was a higher response rate among pharmacists (67%, 27/40) than among physicians (30%, 30/100). Following screening, three questionnaires were excluded from the physicians’ responses. Total valid responses were, therefore, 54, comprising an equal number (27) of physicians and pharmacists.

Table 1 presents the results for the survey (Supplement). (Detailed demographic information is attached as Appendix 2 in Supplemental material).

### AMR: prevalence and solutions

Participants were aware of a high AMR prevalence rate, if understudied/underreported. Almost all (96%, 52/57) reported AMR to be a problem in Yemen. The majority (56%, 15/27 of physicians and 70%, 17/27 of pharmacists) perceived AMR to be an understudied or underreported problem.

More pharmacists perceived a higher prevalence of AMR in IDPs relative to the general population (63%, 17/27) compared to physicians (48%, 13/27).

Physicians and pharmacists differed in their ranked perception of solutions to the high burden of AMR in Yemen. Physicians were more (37%, 10/27) in support of the enforcement of prescription laws – sale of antimicrobials without prescription – and increased training (30%, 8/27) than in increasing awareness among clients (22%, 6/27). On the other hand, pharmacists were more in support of increased training (59%, 16/27) and increased awareness among clients (22%, 6/27), than enforcement of prescription laws (15%, 4/27).

Interestingly, both ranked the use of AST the lowest – only 11% (3/27) of physicians and 4% (1/27) of pharmacists reported increased testing for susceptibility as an approach to tackling AMR.

### Antibiotic utilization – prescribing/dispensing practices and major antibiotics

Most physicians (74%, 20/27) reported that they felt under pressure to prescribe broad-spectrum versus narrow-spectrum antibiotics.

There were multiple reasons for empirical treatment with antibiotics with the major ones being cost (59%, 16/27) and symptoms of patient (56%, 15/27); and for antibiotic choice, with availability, and broad-spectrum nature (52%, 14/27), respectively being the main reasons. Other reasons for empirical treatment and antibiotic choice were identification (from signs and symptoms) of bacterial infection (41%, 11/27) and convenience (11%, 3/27).

The practice of selling antibiotic without a prescription was reported by most pharmacists (67%, 18/27) as with counter-prescribing (63%, 17/27). The conditions for which antibiotics were demanded or counter-prescribed were: inflammation affecting the throat/mouth or ear (37%, 10/27), followed by infections – Urinary Tract Infections (UTIs) and Respiratory Tract Infections (RTIs) (33%, 9/27, each) and fever with other symptoms (11%, 3/27).

The most important consideration for dispensing of antibiotics by pharmacists were: disease (62%, 16/27); price patient can afford (22%, 6/27) and cost (15%, 4/27).

Penicillins were the most commonly-prescribed and dispensed antibiotic group, followed by quinolones, macrolides and tetracyclines. Of the penicillins, amoxicillin (and amoxicillin/clavulanic acid) were the most commonly prescribed (63%, 17/27) by physicians or dispensed (82%, 22/27) by pharmacists. Ciprofloxacin was the most common quinolone antibiotic prescribed (52%, 14/27) or dispensed (74%, 20/27). There was more community dispensing of macrolides, with pharmacies dispensing more of erythromycin (26%, 7/27) than the healthcare facilities (19%, 5/27), and with azithromycin dispensed only by pharmacists (15%, 4/27). Similarly, the dispensing of tetracyclines was reported only in pharmacies (15%, 4/27).

By comparison, cephalosporins were not as commonly prescribed or dispensed. Physicians reported 22% (6/27) of prescribed ceftriaxone – similar to pharmacists, and 11% (3/27) for other cephalosporins compared to 4% (1/27) pharmacists.

### Antimicrobial stewardship

In terms of capacity to perform AST, two-thirds, 67% (18/27), reported the possibility of performing AST at their facilities: most (56%, 10/18) could do so on site. In the majority of cases (81%, 22/27), however, AST was not performed to inform choice of antibiotics.

The major barriers to the routine use of AST were cost, availability and waiting time for AST results. The average cost of one AST in Yemen is estimated at between 3,500 – 5,000 Yemeni Rial (Key informant), equivalent to USD14-20.^39^ Even allowing for a lower 6-month average exchange rate of 600 Rial to 1 USD, the cost is USD6-9. At a daily wage of USD3.39 in 2017, amidst delayed salary payments and inflation, the cost of one AST is prohibitive for three-thirds of the population who live below the poverty line (USD1.9) in 2020.^40–42^ This is in addition to poor access to laboratory facilities and equipment (Key informant, 2020).

Prescribing guides were available only in 30% (8/27) of health workers’ practices, though most participants, 70% (19/27), reported that their facility had a Medicines Formulary. Less than one-fifth, 19% (5/27), reported that their facilities had a DTC. There is no antibiotic policy for Yemen (Key informant, 2020).

The key findings from the survey are summarized in Figure 1.

**Figure 1.**
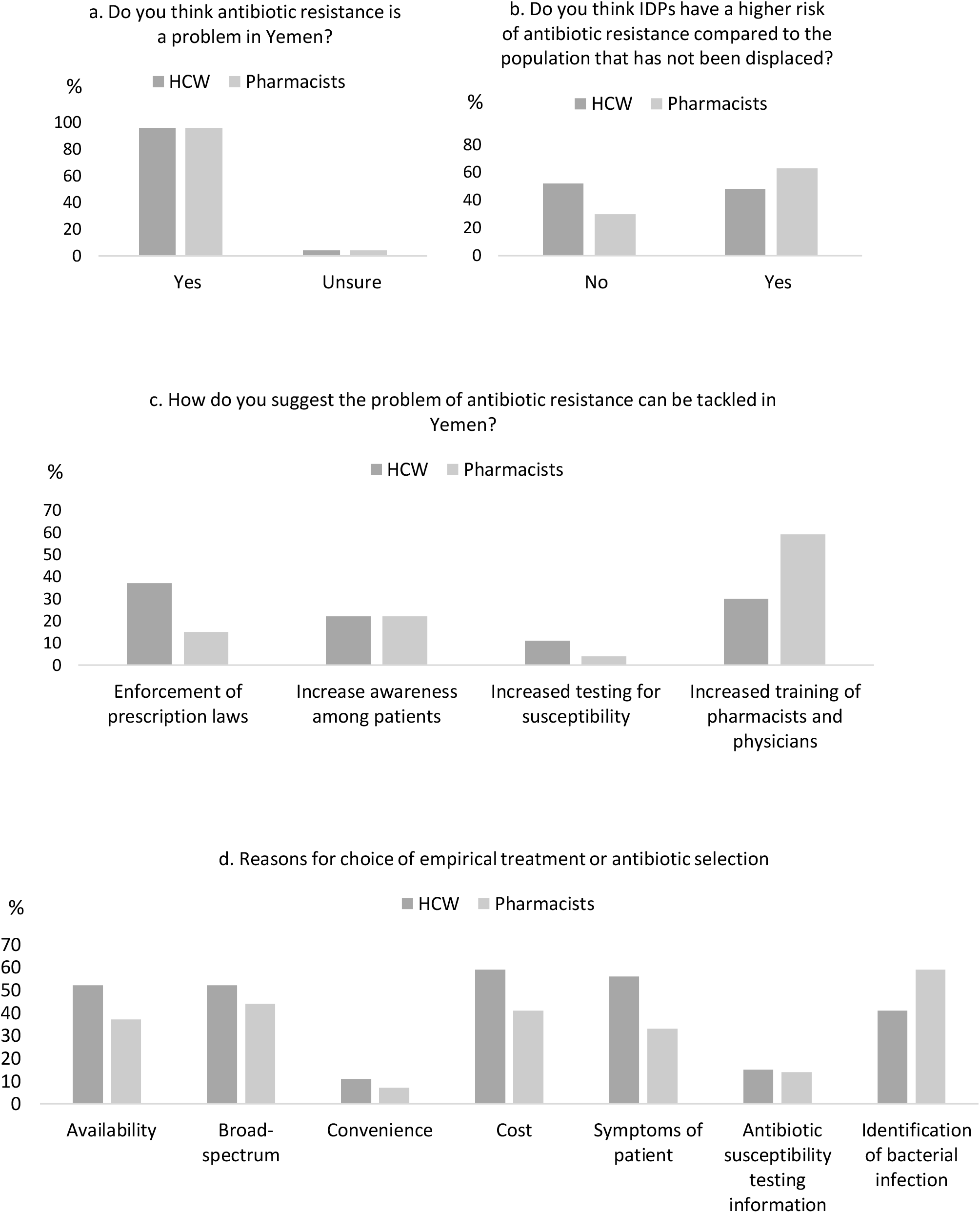
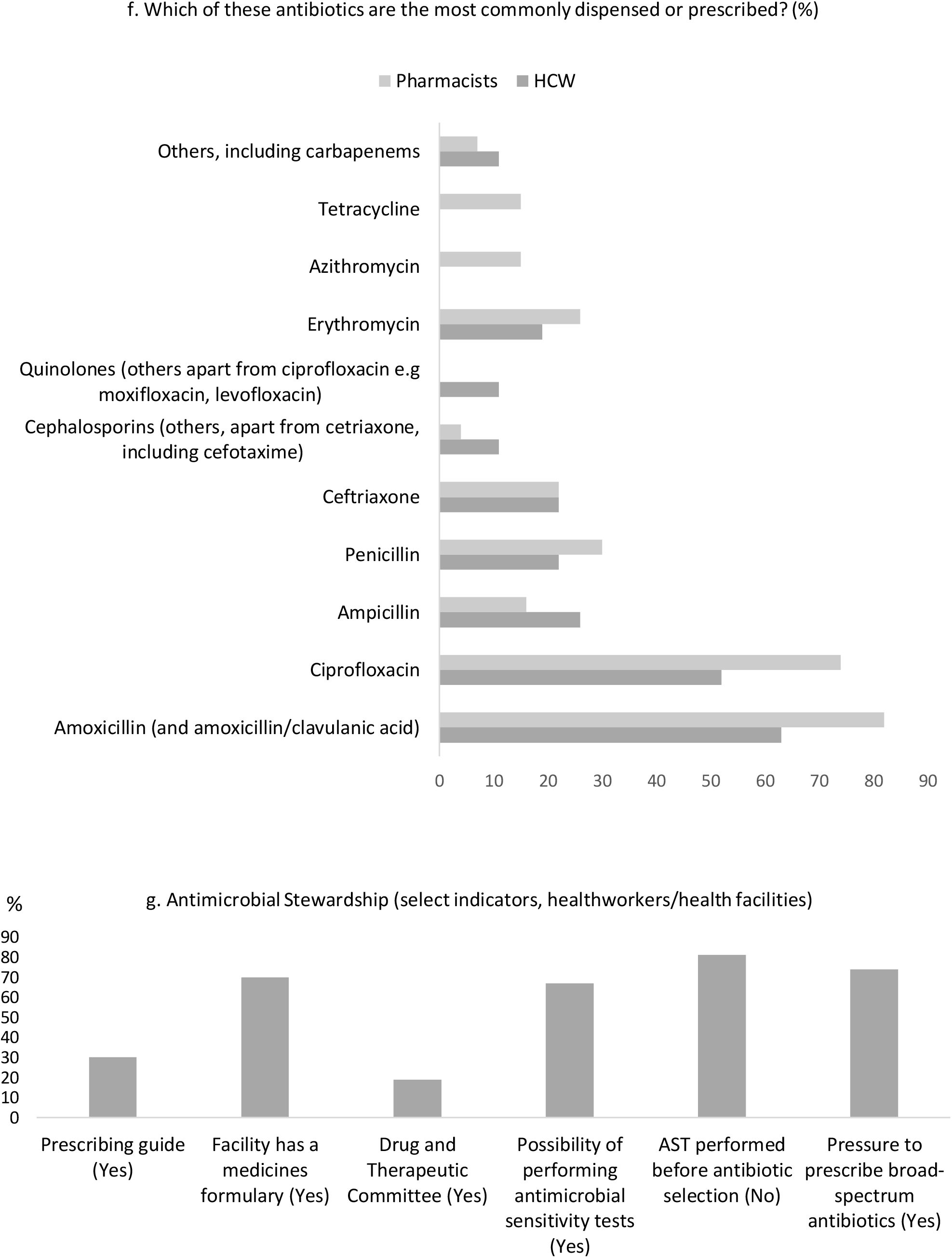
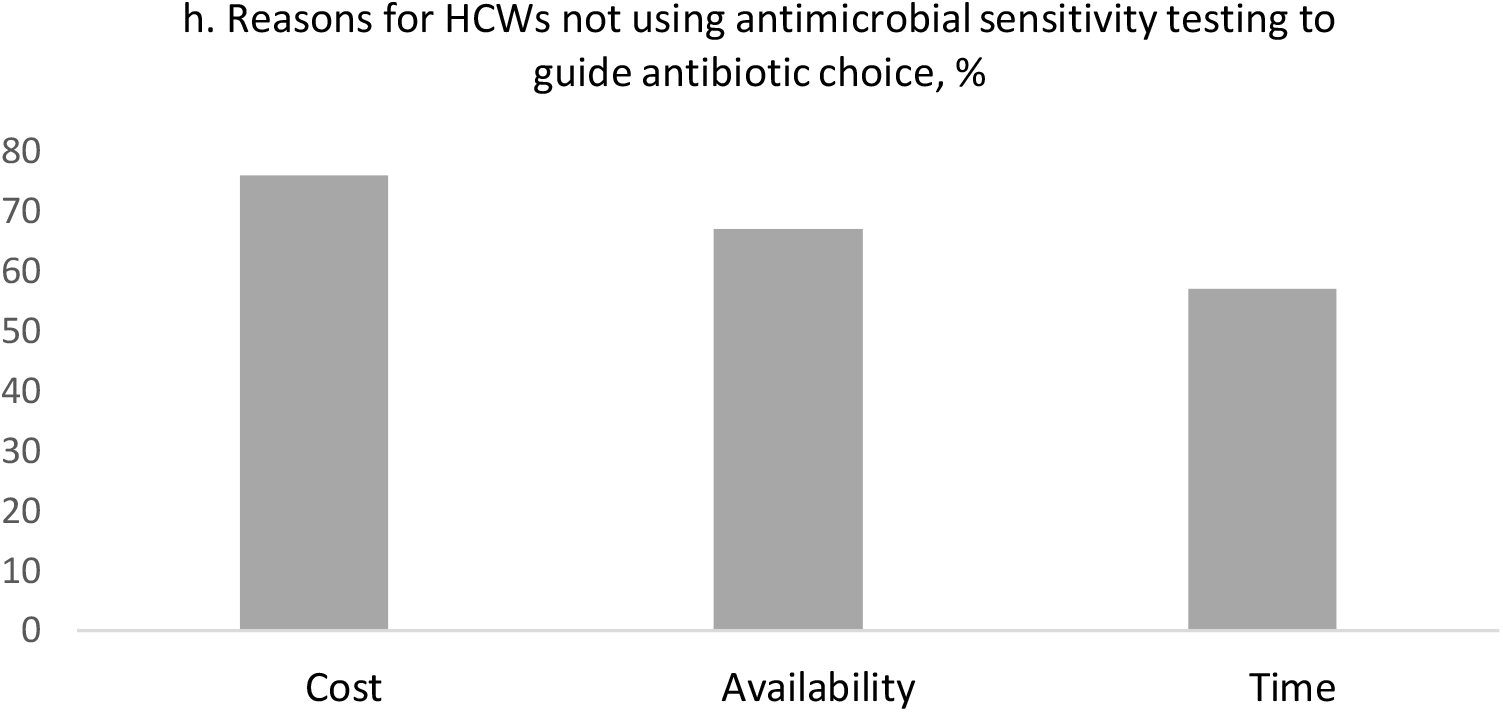
Summary of select results from a survey assessing antimicrobial resistance, use and stewardship in healthcare workers (HCW) and pharmacists in Yemen. a-c. Antimicrobial Resistance (AMR): The majority of respondents (96%) thought AMR to be a problem in Yemen (a); but were more split in terms of if IDPs had the greater risk (b). Among solution options to tackling AMR, both respondents ranked the use of increased antimicrobial sensitivity testing the lowest and ranked increased training of pharmacists and physicians (HCW) the highest. d-f. Antimicrobial Use: There were several overlapping reasons for choice of empirical treatment or empirical antibiotic selection both in prescribing physicians and dispensing pharmacists with cost (price patient can afford), signs and symptoms (including “identification of bacterial infection”) of patient and availability with a preference for broad spectrum antibiotics. In terms of antibacterial agents, amoxicillin (including amoxicillin/clavulanic acid) was the most commonly prescribed (63%, n=27) or dispensed (82%, n=27) followed by ciprofloxacin (52%, n=27, prescribed by HCW; 74%, n=27, dispensed by pharmacists). g-h. Antimicrobial Stewardship: Among select criteria, physicians reported to be under pressure to prescribe broad-spectrum antibiotics (74%, n=27), and even where there was the possibility to perform antimicrobial sensitivity tests (AST) in 67% (n=27) of facilities, these were not used to guide prescribing in the majority (81%, n=27) of cases. The reason for this was largely (74%, n=27) cost (h).

## Discussion

AMR is correlated with an increase in the consumption of antimicrobials.^43–46^ The containment of AMR requires a complete understanding of all factors promoting misuse. This mixed-methods study provides current knowledge of individual-level (awareness and practices) as well as contextual factors related to AMR and stewardship in Yemen – a low-income country in conflict since 2015. There are three salient findings:

- Awareness of a high AMR prevalence rate,
- Inappropriate use of antibiotics, and
- Overall sub-optimal AMS activities, including low prioritization and cost of AST.

### AMR prevalence: perception and solutions

The high level of AMR in Yemen perceived by participants indicates awareness among physicians and pharmacists, considered frontline health workers, in this survey and is corroborated by the earlier mentioned studies reporting laboratory investigations.^24–30^

In Yemen, IDPs may not necessarily be distinguished from the non-displaced. Since 2020, displacement has been more marked among four, out of a total of 22 governates (*Marib, Al Hudaydah, Al Dhale’e, Taizz*, and *Al Jawf*) – mostly in the north – with the greater proportion moving in with relatives or other non-family hosts, rather than into any settlement camps.^47^ IDPs, as they experience the same conditions as their hosts, would, thus, be hypothesized, or expected, to have the same risks, or prevalence, of AMR as the general population, as the results also seem to suggest.

The difference in the ranking by physicians and pharmacists of solutions to the perceived high burden of AMR in Yemen may reflect differential clinical training and practice environments as well as highlight training needs or gaps.^48,49^ In this, our findings are in line with a previous 2016 study among healthcare workers (physicians, pharmacists and nurses) assessing awareness of AMR, possible contributory factors, as well as solutions.^49^

Interestingly, despite the level of awareness, both professionals surveyed in our study ranked the use of AST as the least important solution to the problem of AMR in Yemen.

### Antimicrobial utilization

#### Prescription and dispensing practices

Antibiotic prescribing is a complex activity heavily influenced by contextual individual, social and policy factors, creating ethical dilemmas.^50^ Physicians may feel obligated to prescribe antibiotics both in response to direct patients’ demand and, on the need, to induce rapid relief of symptoms. Both of these, as shown in a systematic review by Rodrigues et al (2013) on physician prescribing behavior, can create a state of tension or fear – one of several factors leading to irrational prescribing.^51^ While the extent to which pressure to prescribe broad-spectrum antibiotics for symptom relief may be related to actual need, for instance infected war-wounds, was not investigated, patients’ demand for antibiotics when not clinically indicated is well documented in the literature.

Failure to prescribe antibiotics when demanded could lead the patient to other physicians willing to do so, or to pharmacists. Hence, this culture is perpetuated. This is common in many geographical regions. As one approach to reduce patient demand, the United Kingdom (UK) mounted a patient awareness campaign to create awareness among the lay population that not all colds require an antibiotic as well as institute stricter prescribing guidelines such that General Practitioners would not prescribe an antibiotic for acute upper RTIs, including cough, common cold, acute pharyngitis, tonsillitis and otitis media, except for the systematically very unwell or with high risk of complications.^52,53^ Generally, educational interventions aimed at creating awareness among clinicians and the public alike have been shown to reduce irrational prescribing, especially for RTIs.^54^

In resource-limited settings, affordability – the price a patient can pay for their medicines – also affects prescribing pattern. This is the case in this study, where physicians reported cost as a major factor both in the choice of antibiotics and in empirical treatment, creating another layer of complexity.

There was pharmacy sale and counter-prescribing of antimicrobials, in common with many others countries especially in the global south, and as previously documented for Yemen.^55,56^ In these contexts, pharmacies are usually the first port-of-call for clients with minor ailments, and, thus, serve an important public health function. Our results conform with a 2014 study by Belkina et al in which 78.2% (n=400) teachers indicated having obtained a non-prescribed antibiotic.^8^ The WHO GAP, as one step to containing AMR, recommends the prescription-only sale of antimicrobials in pharmacies and medicine outlets. However, in the global north, the role of the pharmacist is evolving to include the prescription of medicines, with some evidence of their impact in decreasing irrational prescribing in hospitals.^57^ Enhancing their capacity by equipping private pharmacies in LMICs with rapid diagnostic devices, including the use of mobile-based decision-making algorithms, could leverage this professional capacity in the containment of AMR.^58^ Using these tools, pharmacists can, for example, screen bacterial from viral infections, and advise clients with viral infections including the common cold more appropriately on the choice of medicines, and whether or not antibiotics are required. These tools should also be available at healthcare facilities.^25^

#### Consumption pattern and AMR

Amoxicillin (including amoxicillin/clavulanic acid), a penicillin, was the most commonly prescribed or dispensed antibiotic among the participants. This corresponds with results from several studies assessing antimicrobial utilization in the community in Yemen.^59–61^ In addition, it mirrors the high usage of this antibiotic in the greater Middle Eastern region.^1^ In Yemen, ciprofloxacin is the first choice antibiotic treatment for several infections including UTIs and gastrointestinal infections including typhoid.^24^ Both of these antibiotics were for oral use, with amoxicillin a preferred choice in susceptible RTIs, especially in children, as contained in the national EML and guidelines for child health programs in Yemen which is widely supported by international organizations.

In a recent study of AMR in Aden, Yemen, by Badulla et al (2020), the resistance to amoxicillin/clavulanic acid among seven bacterial species isolated from various clinical specimens – urine, pus and wound – was an average of 65.2%.^24^ This study raises questions about the continued efficacy of amoxicillin and amoxicillin/clavulanic acid in Yemen.

Resistance against ceftriaxone, the most commonly prescribed/dispensed cephalosporin in this survey, was also fairly high at about 62% of microorganisms in their study. In contrast, resistance to ciprofloxacin was relatively low at about 26%.

This study by Badulla et al., while limited to Aden and, therefore, potentially not generalizable, might account in part for the treatment failure incidences reported by participants in this survey. However, a number of other studies conducted in other parts of Yemen note high resistance against antibiotics noted as commonly dispensed by the survey conducted.^25,26,28–30^ For instance, a study from Sana’a found that clinical isolates of *E. coli* collected in 2017 were 96% resistant to amoxicillin-clavulanic acid ^29^ and another study from *Al-Mukalla* found UTI *E. coli* isolates from 2003-2006 were 78.8% resistant to penicillin and 73.1% resistant to cephalosporins.^26^

Other possible reasons for treatment failure may include the use of SF medicines, non-compliance with recommended dosages and the empirical use of broad-spectrum antibiotics. The prevalence of poor-quality medicines is thought to be high ^63^ and have been estimated by various sources to be between 10-60% in Yemen.^64^ As part of its pharmacovigilance, Yemen collects information on suspected therapeutic failure due to poor-quality medicines. Current pharmacovigilance reports on poor-quality antimicrobials put this at <0.012% (Key informant, 2020).

The potential to induce AMR differ according to antibiotic, with the broad-spectrum macrolides and fluoroquinolones being among the most potent inducers (fluoroquinolones can induce AMR even with a single use). The AWaRe classification aims to reduce the use of potent inducers – Watch/Reserve antibiotics – or last choice antibiotics – Reserve – to only serious cases of infection resistant to the Access group of antibiotics. One target of this classification is to increase the use of Access antibiotics, such as penicillins, to 60% by 2050, as one approach to tackling AMR. In this regard, the results of this survey, in almost equal proportions, is both encouraging and concerning.

### Antimicrobial stewardship

Overall, the use of AST to identify resistance and to better guide the prescription process was sub-optimal. While there was capacity to perform AST in the majority of the facilities where the surveyed physicians’ practice, routine use to guide the choice of antimicrobial agent was limited and, in some cases, non-existent. Physicians rather prescribed antibiotics based on clinical and financial considerations, with preference for a broad-spectrum antibiotic. This preference for initial (empirical) treatment with a broad-spectrum antibiotic is possibly informed by the perception of an assumed (pre)existing antibiotic resistance to first-line antibiotics in patients. The irrational prescribing of antibiotics potentially leading to AMR has been described for public hospitals in Yemen, including up to several antibiotics at one time, as also found from this study.^65^

In recognition of the high levels of AMR among certain impacted populations in its health facilities in conflict-affected countries in this region, the MSF routinely use AST including rapid diagnostics and has developed a propriety field device to facilitate AST.^5,66^ However, the challenges in both the development and deployment of these devices, and of AST in certain contexts, must be acknowledged.^67^

Facilitators to increase the uptake of AST might include: increased access to quality-assured AST kits or equipment and consumables including media for both aerobic and aerobic bacteria; and capacity increase and training for laboratory personals, as well as logistical support. Development partners could play a role here in increasing access to AST including rapid diagnostics, through subsidies.

Other structures to ensure AMS were also sub-optimal. Many facilities lacked prescribing guides. The majority had no DTC – an institutional mechanism for ensuring the rational use of medicines, including antibiotics, in a healthcare facility – though most had a Medicines Formulary. These results are in line with a previous survey on AMS in health facilities in Yemen.^68^ Educational interventions to increase the use of AST among professionals, including of AMS in community pharmacies may be needed to address the observed AMS gaps.^69^ Actively promoting AST use in pharmacies as collaborative efforts aimed at AMR containment may also be needed. ^70,71^

Limitation: The small sample size of this study limits its generalizability across the country, as earlier indicated. However, this study advances our understanding of some current challenges in the country with respect to AMR. There is need for more robust studies.

## Conclusion

Active conflicts aggregate multiple social determinants promoting irrational use and AMR. Challenges to AMS in these settings are multifactorial, demanding complete understanding for containment. This study found that while there is awareness of a high AMR prevalence in Yemen among health workers surveyed, there was sub-optimal antimicrobial use practices and AMS. The main barrier to AMS was cost.

There is the need for educational/awareness interventions targeting both providers and consumers of antibiotics to reduce irrational use. Policy changes to the use of amoxicillin as a preferred antibiotic may be required. There is also a strong need to build AMS structures as a priority. These activities, could be built using the WHO antimicrobial stewardship toolkit as a guide.^72^ Barriers to the use of AST could be addressed through the deployment of low-cost AST kits. In all aspects, strong international support would be required.

## Supporting information

Supplement (Table 1; Appendix 1 Questionnaire; Appendix 2 Demographics

## Data Availability

All data included.

## Acknowledgments

This work was made possible by support from the Boston University Social Innovation on Drug Resistance (SIDR) Postdoctoral Program to ESFO and the U.S. Pharmacopeial Convention (USP) Quality Institute to CC.

## Disclosures regarding real or perceived conflicts of interest

None to declare.

