## Supplement (Table 1; Appendix 1 Questionnaire; Appendix 2 Demographics for "Assessing antimicrobial resistance, utilization and stewardship in Yemen: An exploratory mixed-methods study"

*Table 1 Summary of results from the questionnaire survey*

| | | HCW, n=27 (%) | Pharm, n=27 (%) |
|---|---|---|---|
| **I** | **Demographics** | | |
| | *Clients* | | |
| **1** | Children under the age of 12 | | |
| | <10% | 37 | - |
| | Between 10-24% | 22 | - |
| | Between 25-50% | 26 | - |
| | >50% | 15 | - |
| **2** | Adults over the age of 60 | | |
| | <10% | 44 | - |
| | Between 10-24% | 19 | - |
| | Between 25-50% | 33 | - |
| | >50% | 4 | - |
| **3** | Age distribution, average age in years | | |
| | Under 18 | - | 4 |
| | 18-29 | - | 52 |
| | 30-49 | - | 37 |
| | 50-75 | - | 7 |
| **4** | Gender | | |
| | Female | 44 | 26 |
| | Male | 33 | 44 |
| | Prefer not to say | 22 | 30 |

| | | HCW, n=27 (%) | Pharm, n=27 (%) |
|---|---|---|---|
| **5** | IDPs, as proportion of clients | | |
| | <10% | 33 | - |
| | Between 10-24% | 48 | - |
| | Between 25-50% | 7 | - |
| | >50% | 11 | - |
| | *Participants* | | |
| **6** | Profession | | |
| | Health Care Workers | | |
| | Physicians | 96 | - |
| | Others (not stated) | 4 | - |
| | Pharmacists | | |
| | Certified pharmacist | - | 89 |
| | Pharmacy worker | - | 11 |
| **7** | Training (pharmacists) | | |
| | University | - | 67 |
| | Pharmacy | - | 30 |
| | Other | - | 4 |
| **8** | Years of practice | | |
| | <1 | 8 | - |
| | <5 | 44 | 22 |
| | <10 | 30 | 48 |
| | <15 | 19 | 15 |

| | | HCW, n=27 (%) | Pharm, n=27 (%) |
|---|---|---|---|
| **II** | **Perception of AMR** | | |
| **9** | AMR is a problem in Yemen | | |
| | Yes | 96 | 96 |
| | Unsure | 4 | 4 |
| | If yes, in what way? | | |
| | Understudied and/or underreported | 56 (15/27) | 70 (19/27) |
| | High prevalence of AMR | 19 (5/27) | 33 (9/27) |
| | Both understudied/underreported and high prevalence | 19 (5/27) | 11 (3/27) |
| **10** | IDPs have a greater prevalence of AMR | | |
| | No | 52 (14/27) | 30(8/27) |
| | Yes | 48 (13/27) | 63 (17/27) |
| **11** | Treatment failure incidences per month | | |
| | <5 | 22 | 60 |
| | <10 | 44 | 30 |
| | | **HCW, n=27 (%)** | **Pharm, n=27 (%)** |
| | <25 | 26 | 7 |
| | <50 | 4 | - |
| | >50 | 4 | - |
| **12** | Source of information on AMR | | |
| | Customers | - | 37 (10/27) |
| | Friends | - | 4 (1/27) |
| | Social media | - | 4 (1/27) |
| | Training | - | 44 (12/27) |

| | | HCW, n=27 (%) | Pharm, n=27 (%) |
|---|---|---|---|
| **13** | **Tackling AMR** | | |
| | Enforcement of prescription laws | 37 | 15 |
| | Increase awareness among patients | 22 | 22 |
| | Increased testing for susceptibility | 11 | 4 |
| | Increased training of pharmacists and physicians | 30 | 59 |
| | **Antimicrobial stewardship** | | |
| **14** | Prescribing guide (Yes) | 30 | - |
| | If yes, what is the guide? (Local or Foreign, including online) | Foreign | - |
| **15** | Facility has a medicines formulary (Yes) | 70 | - |
| **16** | Dispensing guide | | |
| | Yemen EML | | 0 |
| | Other texts - including WHO, BNF, PNF | | 30 (8/27) |
| | Empirical (based on experience, or studies) | | 22 (6/27) |
| **17** | Drug and Therapeutic Committee (Yes) | 19 | - |
| **18** | Possibility of performing antimicrobial sensitivity tests (Yes) | 67 (18/27) | - |
| | If yes (on-site) | 56 (10/18) | - |
| **19** | AST performed before antibiotic selection (No) | 81 | - |
| | a. If not, reason for not performing: | | |
| | Cost | 76 (16/21) | - |
| | Availability | 67 (14/21) | - |
| | Time | 57 (12/21) | - |
| | b. If yes, how often is it used as a proportion of all cases | | - |
| | <10 | 50 (6/12) | - |
| | Between 10-24 | 17 (2/12) | - |
| | Between 25-49 | 17 (2/12) | - |

| | | HCW, n=27 (%) | Pharm, n=27 (%) |
|---|---|---|---|
| | 75-99 | 17 (2/12) | - |
| **III** | **Practices** | | |
| **20** | Pressure to prescribe broad-spectrum antibiotics (Yes) | 74 (20/27) | |
| **21** | Reasons for choice of empirical treatment or antibiotic selection | | |
| | Availability | 52 (14/27) | 37 (10/27) |
| | Broad-spectrum | 52 (14/27) | 44 (12/27) |
| | Convenience | 11 (3/27) | 7 (2/27) |
| | Cost | 59 (16/27) | 41 (11/27) |
| | Symptoms of patient | 56 (15/27) | 33 (9/27) |
| | Antibiotic susceptibility testing information | 15 (4/27) | 14 (4/27) |
| | Identification of bacterial infection | 41 (11/27) | 59 (16/27) |
| **22** | Antibiotics most commonly prescribed or dispensed | | |
| | a. Spectrum | | |
| | Broad-spectrum | - | 78 |
| | Narrow-spectrum | - | 11 |
| | Both equally | - | 11 |
| | b. Individual agents/ groups | | |
| | Penicillin | | |
| | Amoxicillin (and amoxicillin/clavulanic acid) | 63 (17/27) | 82 (22/27) |
| | Ampicillin | 26 (7/27) | 16 95/27) |
| | Penicillin | 22 (6/27) | 30 (8/27) |
| | Cephalosporins | | |
| | Ceftriaxone | 22 (6/27) | 22 (6/27) |
| | Cephalosporins (others, apart from ceftriaxone, including cefotaxime) | 11 (3/27) | 4 (1/27) |
| | Fluoroquinolones | | |

| | | HCW, n=27 (%) | Pharm, n=27 (%) |
|---|---|---|---|
| | Ciprofloxacin | 52 (14/27) | 74 (20/27) |
| | Quinolones (others apart from ciprofloxacin e.g., moxifloxacin, levofloxacin) | 11 (3/27) | - |
| | Macrolides | | |
| | Erythromycin | 19 (5/27) | 26 (7/27) |
| | Azithromycin | - | 15 (4/27) |
| | Tetracyclines | | |
| | Tetracycline | - | 15 (4/27) |
| | Others, including carbapenems | 11 (3/27) | 7 (2/27) |
| **23** | Sale without prescriptions (Yes) | - | 67 |
| **24** | Counter-prescribing (Yes) | - | 63 |
| **25** | Conditions for which antibiotics demanded | | |
| | Fever with other symptoms | - | 11 (3/27) |
| | Inflammation, including pharyngitis, tonsilitis, otitis media | - | 37 (10/27) |
| | Infection - UTI, RTI, including both upper and lower | - | 33 (9/27) |
| **26** | Most important consideration for dispensing/counter-prescribing | | |
| | Cost | - | 15 (4/27) |
| | Disease | - | 62 (16/27) |
| | Price patient can afford | - | 22 (6/27) |
| **27** | Daily antibiotic prescribing incidence | | |
| | <5 | 7 | 26 |
| | <10 | 22 | 22 |
| | <25 | 48 | 37 |
| | <50 | 19 | 7 |
| | >50 | 4 | 7 |
| **28** | Daily antibiotic prescriptions received | | |

| | HCW, n=27 (%) | Pharm, n=27 (%) |
|---|---|---|
| <5 | - | 15 |
| <10 | - | 41 |
| <25 | - | 41 |
| ≥50 | - | 8 |

**Appendix 1 Questionnaires**

<u>A. Healthcare workers</u>

1-Among the patients you see, approximately what percentage are children under 12?

- less than 10%
- between 10-24%
- between 25-50%
- Over 50%

2-Among the patients you see, approximately what percentage are over 60?

- less than 10%
- between 10-24%
- between 25-50%
- Over 50%

3-Do you see more male or female patients?

- Male
- Female
- Prefer not to say

4-Among the patients you see, approximately what percentage of patients that you see are IDPs?

- less than 10%
- between 10-24%
- between 25-50%
- Over 50%

5-Do you think antibiotic resistance is a problem in Yemen?

- Yes
- No

- Unsure

5A. If yes, in what way? (Check all that apply)

- Understudied problem
- Underreported problem
- High levels of AMR are prevalent

6-Do you think IDPs have a higher risk of antibiotic resistance than population that has not been displaced?

- Yes
- No
-

7-In your practice, is there a guide to prescribing antibiotics?

- Yes
- No

7A. If yes above, please describe the guide to prescribing antibiotics ___________________

8-Does the facility where you work have a Drug and Therapeutic Committee responsible for rational use of medicines?

- Yes
- No

9-Do you use Medicine formulary for your work in the health facility?

- Yes
- No

10-Is it possible to perform laboratory antibiotic susceptibility testing on-site or off-site?

- Yes
- No

10A. If yes, is it on-site or off-site?

- On-site
- Off-site

11-How often are antibiotics prescribed a day?

- less than 5 incidences
- less than 10 incidences
- less than 25 incidences
- less than 50 incidences
- 50 or more incidences

12-How often do you see antibiotic treatment failure within a month?

- less than 5 incidences
- less than 10 incidences
- less than 25 incidences
- less than 50 incidences
- 50 or more incidences

13-Are antibiotic susceptibility tests of bacteria performed before antibiotic selection?

- Yes
- No

13A. If not, why? (Check all that apply)

- Cost
- Availability
- Time

13B. If Yes, how often is it used?

- less than 10% of cases
- 10-24% of cases
- 25-49% of cases
- 50-74% of cases
- 75-99% of cases
- Always (100%)

14- Which out of these antibiotics are most commonly prescribed? (Please choose up to three and specify for other)

- Tetracycline
- Ciprofloxacin
- Erythromycin
- Amoxicillin
- Ampicillin
- Penicillin
- Other: ____________________________

14A. If other, please list most commonly prescribed antibiotics_________________________

15-Why do you choose a particular antibiotic? (Check all that apply and specify for other)

- Cost
- Availability
- Identification of bacterial infection
- Antibiotic susceptibility testing information
- Convenience
- Broad-spectrum
- Symptoms of patient
- Other: ____________________________

16- Do you feel pressured to prescribe broad-spectrum (tetracycline, ciprofloxacin, erythromycin, amoxicillin, ampicillin) antibiotics vs narrow-spectrum antibiotics (penicillins)?

- Yes
- No

17- How do you suggest the problem of antibiotic resistance can be tackled in Yemen? (Choose one)

- Increase testing for susceptibility
- Increase awareness among patients
- Enforce prescription laws
- Improved training of pharmacists and doctors
- Other: ____________________________

18-What is your profession? (e.g., doctor, nurse, healthcare worker, etc.).

- Doctor
- Nurse
- Healthcare worker
- Other: ____________________________

19- How many years of professional experience do you have?

- less than 1 year
- less than 5 years
- less than 10 years
- less than 15 years
- 15 years or more

**B. Pharmacists**

1-Do you think antibiotic resistance is a problem in Yemen?

- Yes
- No
- Unsure

1A. If yes, in what way? (Check all that apply)

- Understudied problem
- Underreported problem
- High levels of AMR are prevalent

2-Do you think IDPs have a higher risk of antibiotic resistance compared to the population that has not been displaced?

- Yes
- No

3-How did you hear about antibiotic resistance?

- Friends
- Family
- Customers
- Training
- Social Media

4-Do you sell over the counter antibiotics?

- Yes
- No

5-How often are prescriptions received for antibiotics a day?

- less than 5 incidences
- less than 10 incidences

- less than 25 incidences
- less than 50 incidences
- 50 or more incidences

6-How often are antibiotics dispensed a day?

- less than 5 incidences
- less than 10 incidences
- less than 25 incidences
- less than 50 incidences
- 50 or more incidences

7-How often do patients return due to treatment failure within a month?

- less than 5 incidences
- less than 10 incidences
- less than 25 incidences
- less than 50 incidences
- 50 or more incidences

8-Which antibiotics are most commonly dispensed? (Please choose up to three)

- Tetracycline
- Ciprofloxacin
- Erythromycin
- Amoxicillin
- Ampicillin
- Penicillin
- Other: ____________________________

If other, please list most commonly dispensed antibiotics_____________________________

9-What conditions do you see the highest demand for antibiotics? ____________________

10-What is the average age of customers?

- Under 18
- 18-29
- 30-49
- 50-75
- Over 75

11-Are customers more often male or female?

- Male
- Female
- Prefer not to say

12-Do you counter-prescribe antibiotics?

- Yes
- No

13-What guide do you use for counter-prescribing antibiotics? ________________________

14-Which antibiotics are most commonly counter-prescribed?

- Tetracycline
- Ciprofloxacin
- Erythromycin
- Amoxicillin
- Ampicillin
- Penicillin
- Other: ___________________________

If other, please list most commonly counter-prescribed antibiotics_____________________

15-What is the rationale for empiric antibiotic selection? (Check all that apply)

- Cost
- Availability
- Identification of bacterial infection
- Antibiotic susceptibility testing information
- Convenience
- Broad-spectrum
- Symptoms of patient
- Other: ____________________________

16-What is the most important consideration for the dispensing/counter-prescribing of antibiotics?

- Cost
- Price patient can pay
- Availability of antibiotics
- Disease
- Other: ____________________________

17-Which type of antibiotics are of higher demands; broad spectrum (tetracycline, ciprofloxacin, erythromycin, amoxicillin, ampicillin) or narrow spectrum antibiotics?

- Broad-spectrum
- Narrow-spectrum
- Equal

18-How do you suggest the problem of antibiotic resistance can be tackled in Yemen? (Choose one)

- Increased testing for susceptibility
- Increased training of pharmacists and doctors
- Increase awareness among patients
- Enforcement of prescription laws
- Other: ____________________________

19-What is your profession?

- Certified pharmacists
- Worker in pharmacy

20-What is your professional training?

- University degree
- Training at a pharmacy
- Other: ____________________________

21-What is your profession and years of experience?

- less than 1 year
- less than 5 years
- less than 10 years
- less than 15 years
- 15 years or more

**Appendix 2 Demographics**

Participants: Physicians were almost all physicians, 96% (26/27) (there was no response from one participant). Pharmacists were mostly, 89% (24/27), certified pharmacists, with the rest 11% (3/27), being pharmacy workers. The majority had a University degree, 67% (18/27). 30% (8/27) reported being trained at a pharmacy, with one reporting "other" training.

Clients: Physicians reported seeing a different mix of children under the age of 12 years among clients that use their facilities. Most, 37% (10/27), reported that children comprised <10%, followed by 26% (7/27) who reported children as comprising between 25-50% of clients and 22% (6/27) where children comprised between 10-24%. In a few facilities, 15% (4/27), children comprised more than 50% of clients. This mix possibly reflecting participants' facility type or specialization.

On the other hand, most pharmacists, 52% (14/27), reported that their clients were likely to be adults between the ages of 18-29 years, followed by 37% (10/27) who reported the age range to be between 30-49 years. Clients under 18 and above 60 years comprised less than 10% respectively.

In terms of gender, when identified, most clients at hospitals were females (44%, 12/27); while most clients at pharmacies were males (44%, 12/27). More than one-fifths of participants chose not to indicate the gender of their clients: 22% (n=27) among physicians and 30% (n=27) for pharmacists.

IDPs as a proportion of clients at healthcare facilities varied, possibly with geographical location (north or south) as well as facility type, with all facilities seeing between less than 10% to more than 50%. Almost half of the health workers, 48% (13/27), reported IDPs to comprise between 10-24% of clients, followed by 33% (9/27) reporting this as less than 10%. An almost equal proportion saw either between 25-50%, or more than 50%, respectively 7% (2/27) and 11% (3/27).
